# Stochastic forecasting of COVID-19 daily new cases across countries with a novel hybrid time series model

**DOI:** 10.1101/2020.10.01.20205021

**Authors:** Arinjita Bhattacharyya, Tanujit Chakraborty, Shesh N. Rai

**Author notes:** Corresponding author’s contact. These authors have contributed equally (Arinjita Bhattacharyya, Tanujit Chakraborty).

## Abstract

An unprecedented outbreak of the novel coronavirus (COVID-19) in the form of peculiar pneumonia has spread globally since its first case in Wuhan province, China, in December 2019. Soon after, the infected cases and mortality increased rapidly. The future of the pandemic’s progress was uncertain, and thus, predicting it became crucial for public health researchers. These future predictions help the effective allocation of health care resources, stockpiling, and help in strategic planning for clinicians, government authorities, and public health policymakers after understanding the extent of the effect. The main objective of this paper is to develop a hybrid forecasting model that can generate real-time out-of-sample forecasts of COVID-19 outbreaks for five profoundly affected countries, namely the USA, Brazil, India, UK, and Canada. A novel hybrid approach based on the Theta method and Autoregressive neural network (ARNN) model, named Theta-ARNN (TARNN) model, is developed. Daily new cases of COVID-19 are nonlinear, non-stationary, and volatile; thus a single specific model cannot be ideal for future prediction of the pandemic. However, the newly introduced hybrid forecasting model with an acceptable prediction error rate can help healthcare and government for effective planning and resource allocation. The proposed method outperforms traditional univariate and hybrid forecasting models for the test data sets on an average.

## 1. Introduction

In December 2019, clusters of pneumonia cases caused by the novel severe acute respiratory syndrome coronavirus 2 (SARS-Cov-2) were identified at Wuhan, Hubei province in China [19,22,54]. Soon after the emergence of the novel beta coronavirus, World Health Organization (WHO) characterized this contagious disease as a pandemic in March 2020 due to its rapid spread within and outside the highly mobile population of China which got further aggravated by the densely populated location of the sea-food market and time of Chinese new year [47]. It resulted in an exponential increase in the incidence rate (IR) and case-fatality rate (CFR) [37]. As of March 16, 2021, a total of 120,512,041 confirmed global cases and 2,665,742 deaths have been reported worldwide [3].

Researchers have faced significant challenges to forecast real-time COVID-19 cases with traditional mathematical, statistical, and machine learning-based forecasting tools [9,14,29,31,40,56,60]. Studies in March 2020 with simple and efficient forecasting methods such as exponential smoothing model predicted cases ten days ahead, with large confidence intervals, that despite the positive bias, had reasonable forecast error [43]. Previously used linear and exponential model forecasts for better preparation regarding hospital beds, ICU admission estimation, resource allocation, emergency funding, and proposing strong containment measures were conducted [17] that projected ICU admissions in Italy for March 20, 2020. ICU admission and mechanical ventilation for critically ill patients reached their peak shattering the health system of Lombardy, Italy, by end of March 2020 [18]. Health-care workers had to go through the immense mental stress left with a formidable choice of prioritizing young and healthy adults over the elderly for allocation of life support, especially unwanted ignoring of those who are extremely unlikely to survive [13,48]. Real estimates of mortality with 14-day delay demonstrated underestimating the COVID-19 outbreak and indicated a grave future with a global CFR of 5.7% [2]. Some of the countries are eventually in a second wave with cases rising in India, and the UK. The uncertain future has several unanswered questions on the daily trajectory of the pandemic, infection rate, and number of deaths. Thus, real-time forecasting with time series models could respond to these debatable quires to reach a statistically validated conjecture in this current health crisis. Some of the impacting leading-edge research concerning real-time projections of COVID-19 confirmed cases, recovered cases, and mortality using statistical, epidemiological and machine learning models are as follows. Short-term forecasting of cumulative confirmed cases was produced using autoregressive integrated moving average (ARIMA), Cubist regression, Random Forest, Ridge, Support Vector Regression (SVR) and Stacking-Ensemble Learning (SEL) model in [46] whereas Exponential smoothing model produced ten days ahead forecasts of actual cases within 90% CI and forecasts reflect the significant increase in the trend of global cases with growing uncertainty [43]. Forecasting and nowcasting domestic and international COVID-19 cases is also done with epidemiological compartmental models, like Susceptible-Exposed-Infectious-Recovered (SEIR) and SIR models [14,45,56]. Modern machine learning techniques like neural networks and black-box deep learning models were also used for COVID-19 forecasting [50,52]. However, forecasting COVID-19 cases is harder and this is primarily due to the following major factors [28]: (a) Very less amount of data is available; (b) Less understanding of the factors that contribute to it; (c) Model accuracy is constrained by our knowledge of the virus. With an emerging disease such as COVID-19, many biologic features of transmission are hard to measure and remain unknown; (d) Another source of uncertainty, affecting all models, is that we don’t know how many people are, or have been, infected; (e) We are certainly missing a substantial number of cases due to virologic testing, so models fitted to confirmed cases are likely to be highly uncertain [21].

Time-series forecasting models take input of historical observations and extrapolate the patterns into the future. The past in no way will continue in the future for this pandemic, so it is challenging to model and produce predictions. There are essentially two general approaches to forecasting a time series: (a) generating forecasts from a single model; and (b) combining forecasts from many models (Hybrid). In classical time series forecasting, the ARIMA model is used predominantly for forecasting linear time series [4], which has a significant strong assumption of linearity in the system and homoscedastic error distribution; typically without sudden jumps and bursts. Individual models such as ARIMA, Wavelet ARIMA (WARIMA)[11], and Theta method [1,26] are inadequate to model such situations. Usage of nonlinear time series techniques in infectious disease modeling has successfully demonstrated the success of Artificial Neural Networks (ANN) and Autoregressive Neural Networks (ARNN) [6,32,34,58]. The idea of hybridizing forecasting models is not a new concept and their empirical applications resulted in superior forecasts to their counterparts. Most recently, some hybrid models combining linear and nonlinear time series models are proposed for COVID-19 [9,16] and performed exceptionally well for predicting these epidemics.

Motivated by this, this study considers the time series data sets of coronavirus confirmed cases that show non-linearity, non-stationary and non-Gaussian patterns, thus making decisions based on a discrete model critical and unreliable. Another difficulty in COVID-19 data from the modeling aspect is the unavailability of sufficient data points, which generates biased predictions and estimates, which can be well maneuvered by neural nets [38]. Most of the relevant studies focused on the outbreak’s short-term and long-term forecasts of reported confirmed cases have a broad range of fluctuations, wide confidence intervals, poorly reported data and model specifics, and highly optimistic predictive performance [57]. Hybridization of two or more models is the most common solution [36] for optimizing forecasting performance, and efficiency with unknown complete data characteristics [30]. The importance of hybrid methodology with a fusion of linear and non-linear forecasting models becomes evident in tackling such dynamic/non-linear time series and its inbuilt time-changing variance, complex autocorrelation structure [9,42]. With the growing need for advanced parsimonious hybrid forecasting methods accompanied by precise accuracy and accurate forecasts, the main objectives of this study are:

1. To propose a simple and computationally efficient hybrid forecasting model which generates out-of-sample forecasts (two months) for five profoundly affected countries (United States of America (USA), Brazil, India, UK, and Canada) with higher accuracy as compared to state-of-the-art forecasting methods..
2. To prove the model’s stationarity and ergodicity properties from a statistical point of view.
3. To discuss the merits and future challenges that needs attention while working with the proposal for epidemiological forecasting. We also recommend policy-making decisions, resource allocation based on these forecasts.

Therefore, this study proposes a novel hybrid Theta Autoregressive neural network model (TARNN) model combining Theta and ARNN models that can capture complex COVID-19 data structures. The proposed TARNN model has easy interpretability, produces robust predictions, and is adaptable to seasonality. An RShiny application is also built for TARNN can help practitioners reproducing and updating the forecasts as further data becomes available.

The rest of the paper is organized as follows: Section 2 describes the formulation of the proposed hybrid TARNN model. The ergodicity and stationarity of the proposed hybrid model are discussed in Section 3. In Section 4, we discuss the country-wise COVID-19 confirmed case data sets and experimental results. The discussions about the results and practical implications are given in Section 5.

## 2. Methodology

We start by discussing the single forecasting models to be used in the hybridization, followed by the detailed formulation of the proposed hybrid Theta-ARNN model.

### 2.1. Theta Method

The ‘Theta method’ or ‘Theta model’ is a univariate time series forecasting technique that performs particularly well in M3 forecasting competition and of interest to fore-casters [1]. The method decomposes the original data into two or more lines, called theta lines, and extrapolates them using forecasting models. Finally, the predictions are combined to obtain the final forecasts. The theta lines can be estimated by simply modifying the ‘curvatures’ of the original time series. This change is obtained from a coefficient, called *θ* coefficient, which is directly applied to the second differences of the time series:

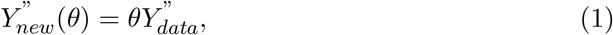

where 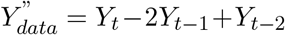 at time *t* for *t* = 3, 4, …, *n* and {*Y*_1_, *Y*_2_, …, *Y*_*n*_} denote the observed univariate time series. In practice, the coefficient *θ* can be considered as a transformation parameter which creates a series of the same mean and slope with that of the original data but having different variances. Now, Eqn. (1) is a second-order difference equation and has solutions of the following form [26]:

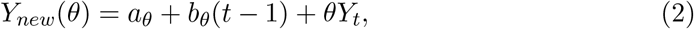

where *a*_*θ*_ and *b*_*θ*_ are constants and *t* = 1, 2, …, *n*. Thus, *Y*_*new*_(*θ*) is equivalent to a linear function of *Y*_*t*_ with a linear trend added. The values of *a*_*θ*_ and *b*_*θ*_ are computed by minimizing the sum of squared differences:

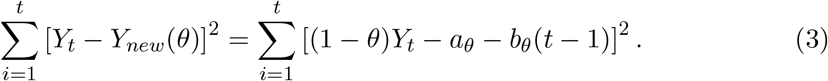

Forecasts from the Theta model are obtained by a weighted average of forecasts of *Y*_*new*_(*θ*) for different values of *θ*. Furthermore, the prediction intervals and likelihood-based estimation of the parameters can be obtained based on a state space model which is demonstrated in [26]. The generalized version of the Theta method is suitable for automatic forecasting of time series [51].

### 2.2. ARNN Model

Artificial Neural Network-based forecasting methods received increasing interest in various applied domains in the late 1990s. A wide variety of neural nets are popularly used for supervised classification, prediction, and nonlinear time series forecasting [59]. The architecture of a simple feedforward neural network can be described as a network of neurons arranged in an input layer, hidden layer, and output layer in a prescribed order. Each layer passes the information to the next layer using weights that are obtained using a learning algorithm [15]. ARNN model is a modification to the simple ANN model especially designed for prediction problems of time series data sets [15]. ARNN model uses a prespecified number of lagged values of the time series as inputs and the number of hidden neurons in its architecture is also fixed [24]. ARNN(*p, k*) model uses *p* lagged inputs of the time series data in a one hidden layered feedforward neural network with *k* hidden units in the hidden layer. Let *<u>x</u>* denotes a *p*-lagged inputs and *f* is a neural network of the following architecture:

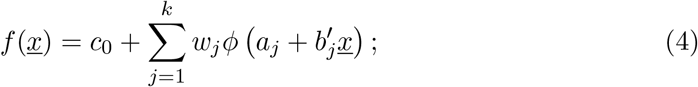

where *c*_0_, *a*_*j*_, *w*_*j*_ are connecting weights, *b*_*j*_ are *p*-dimensional weight vector and *ϕ* is a bounded nonlinear sigmoidal function (e.g., logistic squasher function or tangent hyperbolic activation function). These Weights are trained using a gradient descent backpropagation [49]. Standard ANN faces the dilemma to choose the number of hidden neurons in the hidden layer and the optimal choice is unknown. However, for ARNN model, we adopt the formula *k* = [(*p* + 1)*/*2] for non-seasonal time series data where *p* is the number of lagged inputs in an autoregressive model [24].

### 2.3. Proposed TARNN Model

In this section, we describe the proposed hybrid model based on Theta method and ARNN model and we name it TARNN model. The proposed TARNN model is based on an error remodeling approach and there are broadly two types of error calculations popular in the literature which are given below [35].

#### Definition 2.1.

In the additive error model, the forecaster treats the expert’s estimate as a variable, *Ŷ*_*t*_, and thinks of it as the sum of two terms:

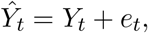

where *Y*_*t*_ is the true value and *e*_*t*_ is the additive error term.

#### Definition 2.2.

In the multiplicative error model, the forecaster treats the expert’s estimate *Ŷ*_*t*_ as the product of two terms:

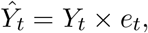

where *Y*_*t*_ is the true value and *e*_*t*_ be the multiplicative error term.

Now, even if the relationship is of product type, in the log-log scale, it becomes additive. Hence, without loss of generality, we may assume the relationship to be additive and expect the errors (additive) of a forecasting model to be random shocks. However, this is violated when there are complex correlation structures in the time series data and less amount of knowledge is available about the data generating process. A simple example is the daily confirmed cases of the COVID-19 cases for various countries where very little is known about the structural properties of the current pandemic. Thus, we need two-stage modeling approach to formulate this complex time series problem. The proposed TARNN model is a hybrid model based on the additive error re-modeling approach. The hybrid TARNN approach consists of three basic steps:

- In the first step of the TARNN model, the Theta method is applied to the time series data to model the linear components of a given time series data set.
- Theta model generates in-sample forecasts and the error series is calculated.
- In the next phase, the residuals (additive errors) generated by the Theta method are remodeled using a nonlinear ARNN model. Finally, both the forecasts obtained from the Theta and ARNN models are combined together to get the final forecasts for the given time series.

The mathematical formulation of the proposed hybrid TARNN model (*Z*_*t*_) is as follows:

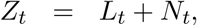

where *L*_*t*_ is the linear part and *N*_*t*_ is the nonlinear part of the hybrid model. We can estimate both *L*_*t*_ and *N*_*t*_ from the available time series data. Let 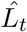 be the forecast value of the Theta model at time *t* and *ϵ*_*t*_ represent the error residuals at time *t*, obtained from the Theta model. Then, we write

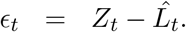

These left-out residuals are further modeled by ARNN model and can be represented as follows:

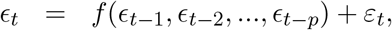

where *f* is a nonlinear function and the modeling is done by the ARNN model as defined in Eqn. (4) and *ε*_*t*_ is supposed to be random shocks. Therefore, the combined forecast can be obtained as follows:

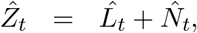

where 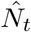 is the forecasted value of the ARNN model. An overall flow diagram of the proposed TARNN model is given in Figure 1.

**Figure 1.**
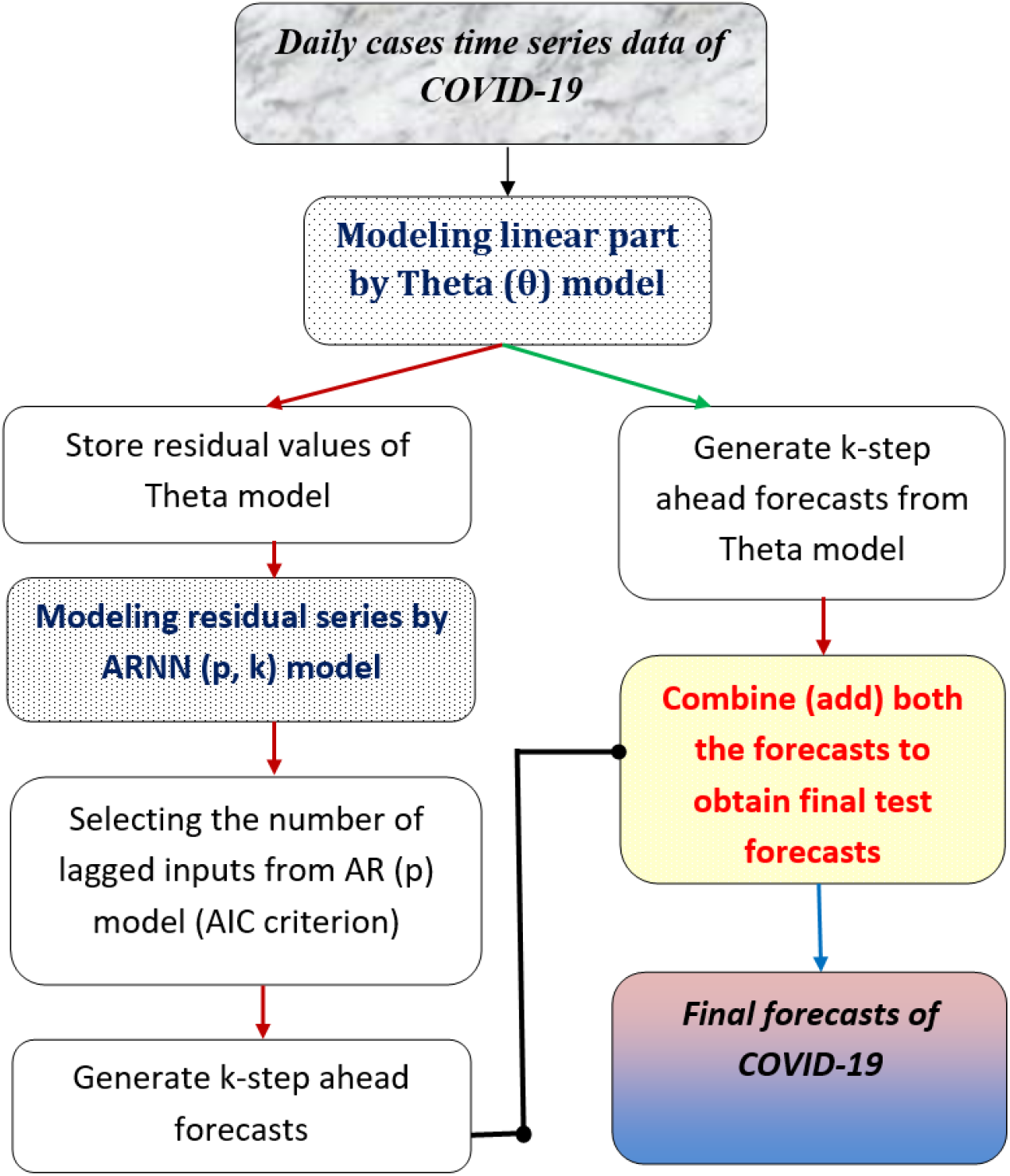
The main workfow of the proposed TARNN model.

In the proposed TARNN model, ARNN is applied to remodel the left-over autocorrelations in the residuals which Theta model individually could not model. Thus, the proposed TARNN model can be considered as an error re-modeling approach. This is important because due to model mis-specification and disturbances in the pandemic time series, the linear Theta model may fail to generate white noise behavior for the forecast residuals. The TARNN approach eventually can improve the predictions for the epidemiological forecasting problem as shown in Section 4.

#### Remark 1.

The idea of additive error modelling is useful for modeling complex time series for which achieving random shocks based on individual forecasting models is difficult. More precisely, the TARNN approach is developed for forecasting the COVID-19 confirmed cases for which the data generating process and the various characteristics of the epidemic are still unknown. The proposed TARNN model only assumes that the linear and nonlinear components of the epidemic time series can be separated individually.

## 3. Ergodicity and Stationarity of the proposed TARNN model

In this section, we derive the results for the ergodicity and asymptotic stationarity of the proposed TARNN model. The ergodicity and stationarity is of particular importance from a statistician’s point of view in time series analysis since for such processes a single realization displays the whole probability of the data generating process. We use several previous results on nonlinear time series and Markov chains to find sufficient conditions for which the overall process is ergodic and stationary [5,7,10,53].

To start, we write the underlying stochastic model of the Theta method using the state space approach. We initialize the model by setting *Y*_1_ = *l*_1_ and then for *t* = 2, 3, … ; and drift term *b*, let

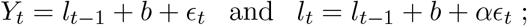

where {ϵ _*t*_} is assumed to follow Gaussian white noise with mean zero and variance *σ*^2^ and *α* is the smoothing parameter for the simple exponential smoothing (SES) model. Now, *Y*_*t*_ follows a state space model which gives forecasts equivalent to SES with drift [26]. Also, *Y*_*t*_ can be written in the following form:

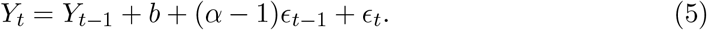

The above is an ARIMA(0,1,1) process with drift term [26]. The left-out residuals of Theta model is further modeled by ARNN process. We consider the ARNN process generated by the additive noise of the ARIMA(0,1,1) process with drift. Let *ϵ*_*t*_ be the process defined by the stochastic difference equation of the following form:

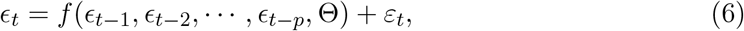

where *ε*_*t*_ is an i.i.d. noise process and *f* (*·*, Θ) is a feedforward neural network with weight (parameter) vector Θ and inputs *ϵ*_*t−*1_, *ϵ*_*t−*2_,…, *ϵ*_*t−p*_. The definition of *f* is given in Eqn. (4).

### 3.1. Time Series as Markov chains

We start by defining the following notations:

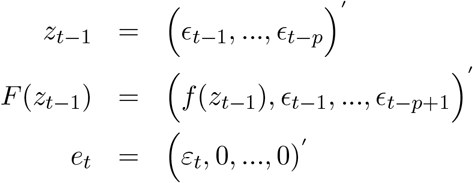

Then Eqn. (6) can be written as follows [10]:

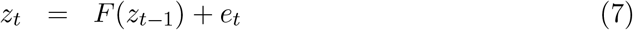

with *z*_*t*_, *e*_*t*_ ∈ ℝ^*p*^. In this section, we show the (strict) stationarity of the state space form, as defined in Eqn. (7). The problem of showing {*z*_*t*_} to be stationary is closely related to the ergodicity of the process [55]. A Markov chain {*z*_*t*_} is called geometrically ergodic if there exists a probability measure *π* on the state space (ℝ^*p*^, 𝔹) and a constant *ρ* > 1 such that

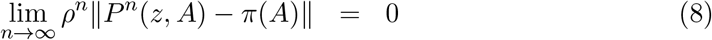

for each *z* ∈ ℝ^*p*^, 𝔹 is the Borel *σ*-algebra on ℝ^*p*^ and ‖ · ‖ denotes the total variation norm. If Eqn. (8) holds for *ρ* = 1, then {*z*_*t*_} is called ergodic.

The definition for *P*^*n*^(*z, A*) can be given as the probability that {*z*_*n*_} moves from *z* to the set *A* ∈ 𝔹 in *n* steps: *P* ^*n*^(*z, A*) = *P* (*z*_*t*+*n*_ ∈*A*|*z*_*t*_ = *z*). Also, expression for *π*(*A*) is as follows:

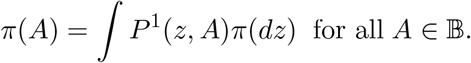

Thus, we call *π* as the stationary measure and the distribution of *z*_*t*_ converges to *π* if {*z*_*t*_} is ergodic. Then, we say {*z*_*t*_} is asymptotically stationary [33]. To establish the ergodicity of TARNN processes, we need the concept of irreducibility and aperiodicity. A Markov chain {*z*_*t*_} is called irreducible if

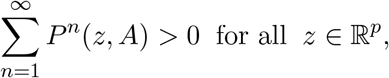

whenever *λ*(*A*) > 0 and *λ* denotes the Lebesgue measure on (ℝ^*p*^, 𝔹). Thus, for an irreducible Markov chain, all parts of the state space can be reached by the Markov chain irrespective of the starting point. Now, an irreducible Markov chain is aperiodic if there exists an *A* ∈ 𝔹 with *λ*(*A*) > 0 and for all *C* ∈ 𝔹, *C ⊆ A* with *λ*(*C*) > 0, there exists a positive integer *n* such that

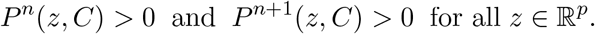

Hence, it is possible that the Markov chain returns to given sets only at specific time points for an aperiodic Markov chain. For most general time series models, irreducibility and aperiodicity cannot be assumed automatically. However, for a TARNN process, these conditions can be checked. In general, it is sufficient to assume the distribution of the noise process to be an absolutely continuous component with respect to Lebesgue measure and the support of the probability density function (PDF) is sufficiently large.

### 3.2. Main Results

It is clear from the above discussion that if the Markov chain is geometrically ergodic, then its distribution will converge to *π* and the corresponding time series will be called asymptotically stationary, see also [53]. Lemma 3.1 states that the state space of the Markov chain cannot be reduced depending on the starting point.

#### Lemma 3.1.

*Let* {*z*_*t*_*} is defined by (7), and let E* |*ε*_*t*_| < ∞ *and the PDF of ε*_*t*_ *is positive everywhere in* ℝ. *Then if f is defined by (4), the Markov chain {z*_*t*_*} is φ-irreducible and aperiodic*.

#### Proof.

Since the support of the PDF of *ε*_*t*_ is the whole real line, that is, the PDF is positive everywhere in ℝ, then we can say that {*z*_*t*_} is *φ;*-irreducible by using [10]. In our case, every non-null *p*-dimensional hypercube can be reached in *p* steps with positive probability (and hence every non-null Borel set *A*). A necessary and sufficient condition for {*z*_*t*_} to be aperiodic is to have a set *A* and positive integer *n* such that *P* ^*n*^(*z, A*) > 0 and *P* ^*n*+1^(*z, A*) > 0 for all *z* ∈*A* [53]. In this case, this is true for all *n* due to consideration of the unbounded additive noise.

The theorem below states the necessary conditions for geometric ergodicity of a Markov chain. This can be obtained using the decomposition technique and ergodicity of stochastic difference equations [7,10].

#### Theorem 3.2.

*Suppose* {*z*_*t*_*} is defined as in (6) and (7), F be a compact set that can be decomposed as F* = *F*_*h*_ + *F*_*d*_, *and the following conditions hold:*

*(i) F*_*h*_(·) *is continuous and homogeneous and F*_*d*_(·) *is bounded;*

*(ii) E*|*ε*_*t*_| < ∞ *and probability distribution function of ε*_*t*_ *is positive everywhere in* ℝ; *then* {*z*_*t*_*} is geometrically ergodic*.

#### Proof.

{*z*_*t*_} satisfies the following equation:

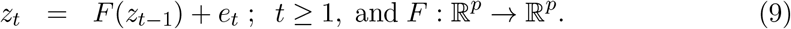

Let *F*_*h*_ be continuous and homogeneous, viz., *F*_*h*_(*cz*) = *cF*_*h*_(*z*) for all *c* > 0, *z*_*t*_ ∈ ℝ^*p*^, and *F*_*d*_ is bounded. Let the origin, *O*, be a fixed point of *F*_*h*_. It is important to note that *ε*_*t*_ satisfies the condition (*ii*) in Theorem 3.2. We are going to show that the existence of a continuous Lyapunov function, *V*, in a neighbourhood of the origin which will ensure the geometric ergodicity of (7).

To start with we let *W ⊆* ℝ^*p*^, the closure of *W* by 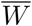 and its boundary by 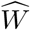. We also let *V* be defined over the closure of the unit ball. We let *p*_0_ = inf_‖*z*‖=1_ *V* (*z*), where ‖ · ‖ denote the Euclidean norm. Also, let *G* be the maximal connected component of 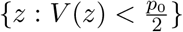 that contains the origin. Then we have 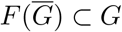.

Let *g*(*z*) = inf {*r* ≥0, *z* ∈ *rG*}, *z* ∈ ℝ^*p*^, where *rG* = {*rz, z* ∈*G*}. Then *g*(*z*) is well defined and it can easily be checked that *g* has the following properties:

1. *g*(*cz*) = *cg*(*z*), for all *c* > 0.
2. There exists 0 < *c* < ℂ < ∞ such that *c* ‖*z*‖ ≤ *g*(*z*) ≤ ℂ‖ *z*‖.
3. 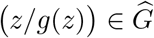
4. There exists *ϵ* > 0, 0 < *θ* < 1 such that for all 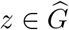, *y* ∈ ℝ^*p*^, we have ‖*y − F* (*z*) ‖ < *ϵ ⇒ y* ∈ *G* and *g*(*y*) < *θ*.

Now, for Eqn. (7), *ε*_*t*_ satisfies *E* |*ε*_*t*_| < ∞. Let *A* ∈𝔹 and *z* ∈ ℝ^*p*^, we define *P* (*z, A*) be the transition probability function as:

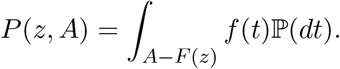

Thus, it holds that

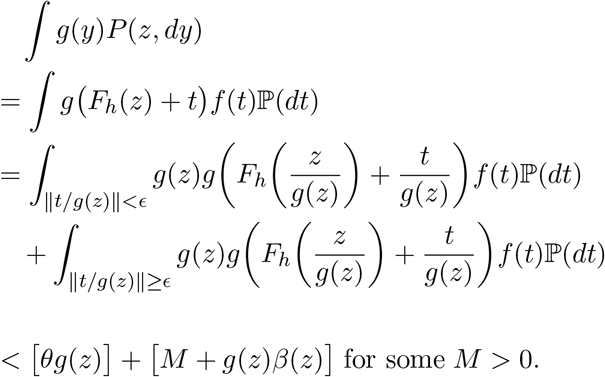

Here |*β*(*z*) | < *B* < ∞ for all *z* and *β*(*z*) → 0 as ‖*z* ‖ *→* ∞. We let *h*(*z*) = *g*(*z*) + 1, and *r* be such that

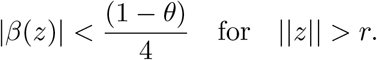

Then for 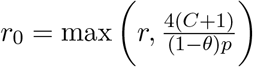, there exists *B*′ such that

1. ∫ *f* (*y*)*P* (*z, dy*) < *B*′ < ∞ when ‖*z*‖ ≤ *r*_0_.
2. 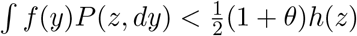 when ‖*z*‖ > *r*_0_.

Using Theorem 4 of [55], we can conclude that {*z*_*t*_} is geometric ergodic.

The next theorem gives the main result for asymptotic stationary of the TARNN model.

#### Theorem 3.3.

*Let E*|*ε*_*t*_| < ∞ *and the PDF of ε*_*t*_ *is positive everywhere in* ℝ, *and* {*ε*_*t*_} and {*z*_*t*_*} are defined as in (6) and (7), respectively. Then if f is a nonlinear neural network as defined in (4), then* {*z*_*t*_} *is geometrically ergodic and* {*ε*_*t*_} *is asymptotically stationary*.

#### Proof.

The noise process *ε*_*t*_ satisfies *E* |*ε*_*t*_| < ∞ by assumption (e.g., Gaussian noise). It is also important to note that neural network activation functions, more precisely logistics or tan-hyperbolic activation functions, are continuous compact functions and have a bounded range. Thus {*z*_*t*_} satisfies all the criteria to be geometrically ergodic and using Theorem (3.2), one can write that for the ARNN process with *F*_*h*_ ≡ 0 and *F*_*d*_ ≡ *F*. Thus, the series {*ε*_*t*_} is asymptotically stationary.

**Remark 2.** Some interpretations and practical implications of the theoretical results are given below:

- The geometric rate of convergence in Theorem 3.3 implies that the memory of TARNN process vanishes exponentially fast. This implies that the simplest version of the proposed model converges to a Wiener process.
- This is important for predictions over larger intervals of time, for example, one might train the network on an available sample and then use the trained network to generate new data with similar properties like the training sample. The results for the asymptotic stationarity can guarantee that the proposed hybrid model can not have growing variance over time.
- From practitioners point of view, when the data is generated by the irreducible TARNN process, the estimated weights are not too far from the true weights. Then, one can draw an indirect conclusion on the statistical nature of the observed data based on the estimated weights.

## 4. Experimental Analysis

Five time series COVID-19 datasets are considered for assessing various forecasting models (individual and hybrid). The datasets are mostly nonlinear, nonstationary, and non-Gaussian in nature. We have used root mean square error (RMSE), mean absolute error (MAE), and mean absolute scaled error (MASE) [27] to evaluate the predictive performance of the models used in this study. Since the number of data points in all the datasets is limited, advanced deep learning techniques will overfit the datasets [20].

### 4.1. Data Description

Data is collected from the “Our World in Data” public repository (Link: https://ourworldindata.org/coronavirus) on five countries: USA, Brazil, India, UK, and Canada that have the leading number of confirmed cases of COVID-19. These are univariate time-series from inception-Feb 26, 2021 of confirmed cases that are analyzed for generating future outbreak predictions. We have taken last 60 data points as test points on which the forecasting models are evaluated. A summary of the COVID-19 data sets of confirmed cases and their descriptions are presented in Table 2. A pictorial view of the training data sets along with auto-correlation function (ACF) and partial ACF (PACF) plots are given in Table 1.

**Table 1.**
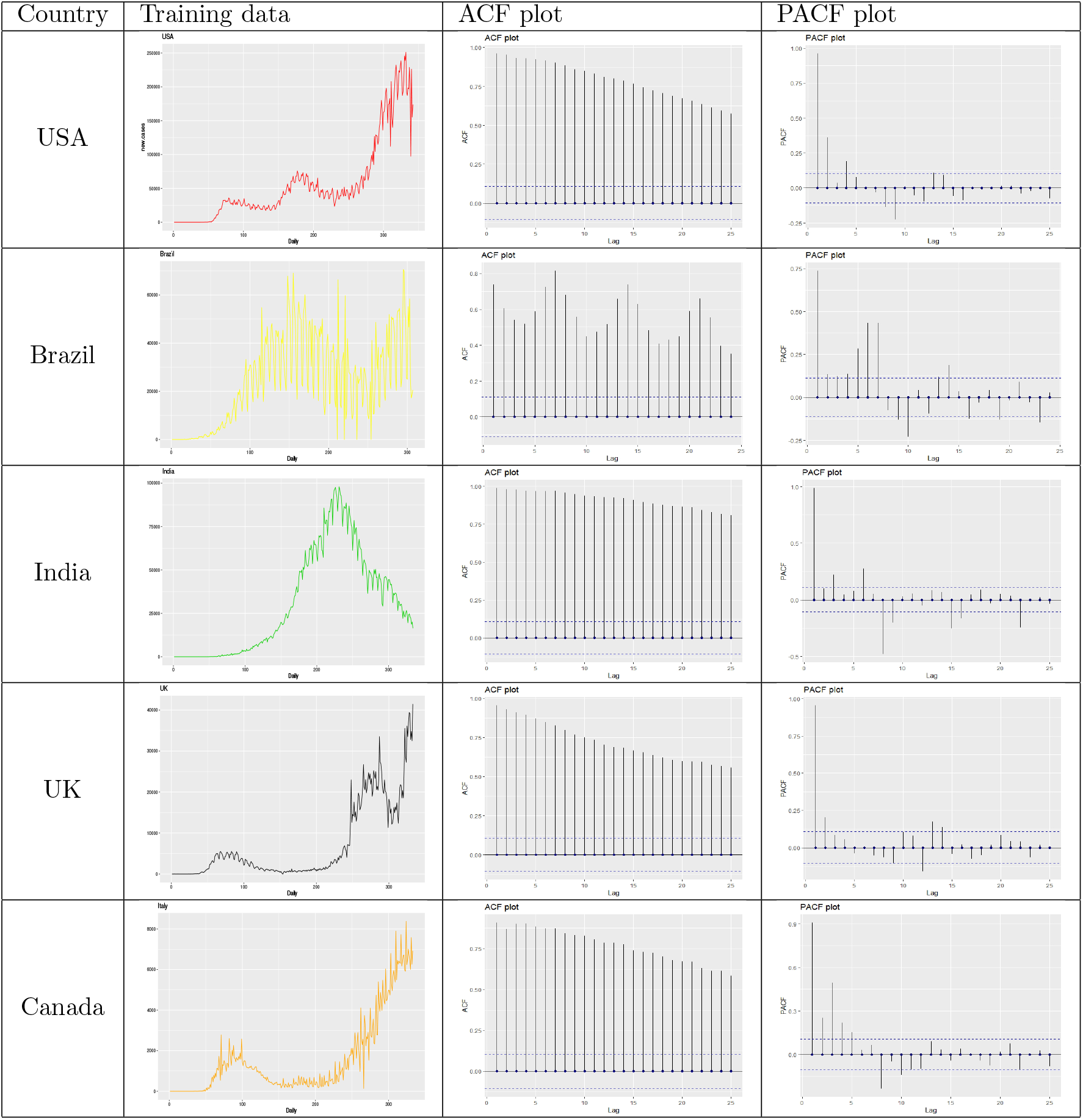
Training data sets (new daily cases) and corresponding ACF, PACF plots for all the countries considered in this study.

**Table 2.** Description of COVID-19 datasets.

| Country Name | Start-End Date | No. of Days | Minimum Value | Maximum Value | Train size | Test size |
| --- | --- | --- | --- | --- | --- | --- |
| USA | 22/01/2020 - 26/02/2021 | 402 | 0 | 251161 | 342 | 60 |
| Brazil | 26/02/2020 - 26/02/2021 | 367 | 0 | 70574 | 307 | 60 |
| India | 30/01/2020 - 26/02/2021 | 394 | 0 | 97894 | 334 | 60 |
| UK | 31/01/2020 - 26/02/2021 | 393 | 0 | 41460 | 333 | 60 |
| Canada | 26/01/2020 - 26/02/2021 | 398 | 0 | 10100 | 338 | 60 |

### 4.2. Performance Evaluation Metrics

The performance of different forecasting models are evaluated based on RMSE, MAE, and MASE metrics for these five time series [24]:

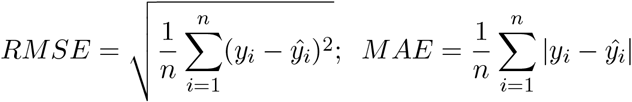

where, *y*_*i*_ is the actual value, *ŷ*_*i*_ is the predicted value, and *n* denotes the number of data points. MASE is computed using ‘mase’ function available under the “Metrics” package in R statistical software. By definition, the lower the value of these performance metrics, the better is the performance of the concerned forecasting model.

### 4.3. Analysis of Results

A schematic diagram is presented in Figure 2 to give an outline of the models to be used in this section. We start the experimental evaluation for all data sets with the classical ARIMA(*p, d, q*) using ‘*forecast* ‘ [25] statistical package in the R statistical software [44]. The nonlinear and nonseasonal time series of these countries were modelled with traditional and advanced methods such as ARIMA, Theta, WARIMA, ETS, TBATS, ARNN, and two popularly used hybrid models, namely hybrid ARIMA-ARNN [8], hybrid ARIMA-WARIMA [9], and the newly introduced hybrid Theta-ARNN (TARNN) models. For the accuracy of prediction, the data was partitioned into training and test sets, where the test set consisted of last 60 data points, and the training set included (inception - 60+1) data points. Table 3 gives the essential details about the functions and packages required for the implementation of standard individual models.

**Table 3.** R functions and packages for standard forecasting model implementations.

| Model | R function | R package | Reference |
| --- | --- | --- | --- |
| ARIMA | auto.arima | forecast | [25] |
| ETS | ets | forecast | [25] |
| TBATS | tbats | forecast | [25] |
| Theta | thetaf | forecast | [25] |
| ARNN | nnetar | forecast | [25] |
| WARIMA | WaveletFittingarma | WaveletArima | [39] |

**Figure 2.**
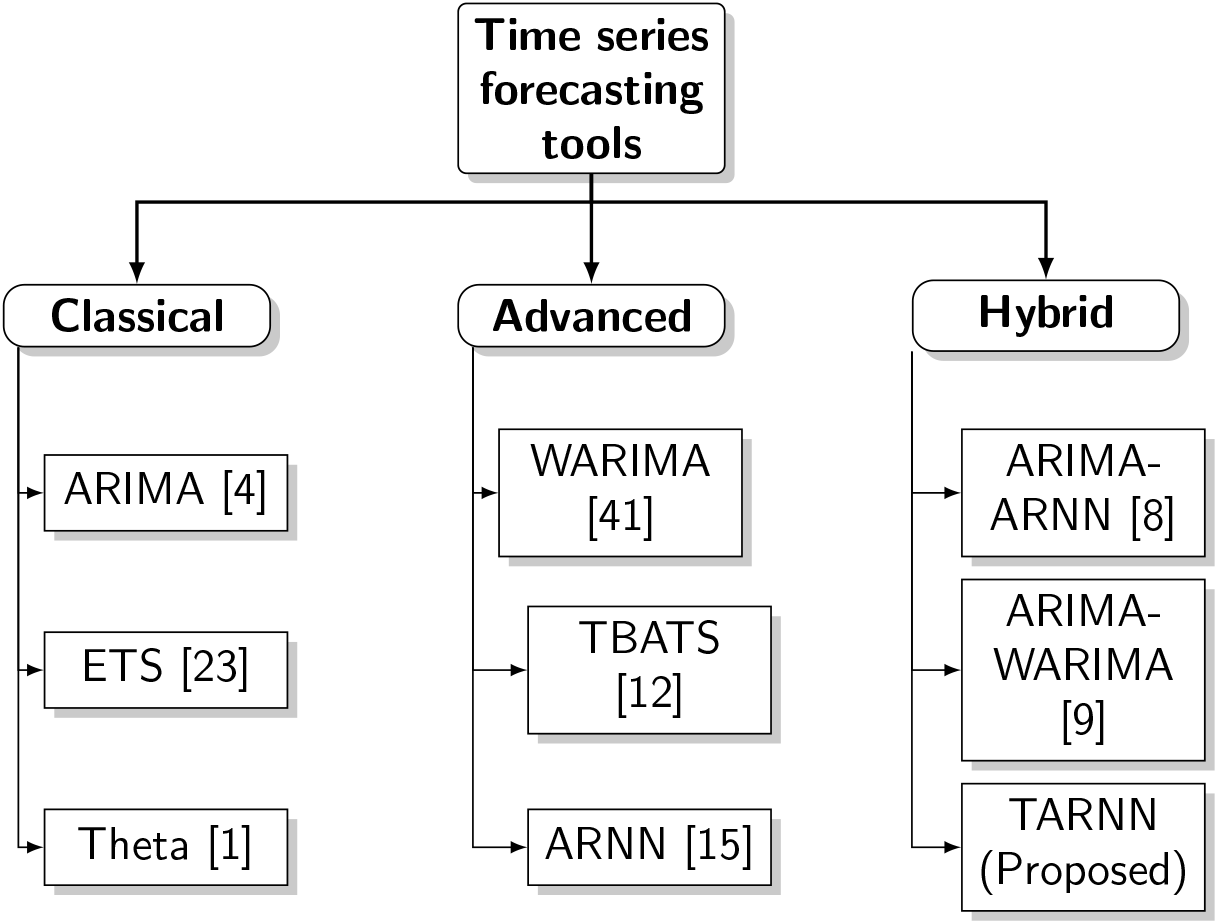
Time series forecasting tools (available and proposed) used in this study.

In the proposed TARNN model, linear modelling is conducted with Theta model using ‘thetaf’ function under the “forecast” package in R statistical software. Nonlinear modelling with ARNN approach is carried out with “caret” package using ‘nnetar’ function in R statistical software. Furthermore, Theta residuals are modelled with ARNN(*p, k*) model having a pre-defined Box-Cox transformation set *λ* = 0 to ensure the forecast values to stay positive. The values of *p* and *k* are obtained by training the network, which is a data-dependent approach as discussed in Section 2.2. Further, both the linear and nonlinear forecasts are summed up to obtain the final forecasts. Theta model was fitted to five data sets namely USA, Brazil, India, UK, and Canada. Theta model residuals for these five countries were trained using ARNN(15,8), ARNN(21,11), ARNN(25,13), ARNN(24,12), ARNN(12,6), and ARNN(9,5) models with an average of 500 networks for all five data sets, each of which is a 15-8-1, 21-11-1, 25-13-1, 24-12-1, 12-6-1, and 9-5-1 networks with 137, 254, 352, 313, 85, and 56 weights and with 5807711, 309816, 20735, 883.6, 209396, and 59477 estimated *σ*^2^, respectively. Finally, the predicted results of both Theta and ARNN models are added together to obtain the estimated forecasts of the proposed TARNN model. Rest of the hybrid models were compared in the similar fashion.

We compared our proposed TARNN model with traditional single models (ARIMA, TBATS, Theta, ARNN, WARIMA, ETS) along with hybrid ARIMA-WARIMA model, hybrid ARIMA-ARNN model and the experimental results are reported in Table 4. The performance of the proposed hybrid Theta-ARNN (TARNN) model is superior compared to all traditional individual and hybrid models on average. The theoretically proven asymptotic stationarity of the proposed hybrid model also suggests that the model cannot have a growing variance over time. The consistency and adequacy in the experimental results empirically approves the same. Thus, the efficacy of the proposed methodology of the proposed hybrid model is experimentally validated. Nevertheless, no model can have dominant advantage and this is related to no free lunch theorem in statistical learning [20]. Forecast of COVID-19 cases for March 20-29, 2021 are generated using proposed TARNN model and shown in Figure 3.

**Table 4.** Quantitative measure of performances of forecasting methods on the COVID-19 test data sets for five countries. Best results are made ‘bold’. Proposed TARNN beats all the models for three out of five data sets and remain competitive for other two data sets.

| Methods | USA | UK | India | Canada | Brazil |
| --- | --- | --- | --- | --- | --- |
| MASE values |  |  |  |  |  |
| ARIMA | 3.55 | 5.53 | 0.67 | 2.08 | 1.55 |
| Theta | 3.24 | 5.88 | 1.89 | 2.08 | 1.31 |
| WARIMA | 4.97 | 6.46 | 5.26 | 1.8 | 1.53 |
| ETS | 2.47 | 5.62 | 1.17 | 1.65 | 1.86 |
| TBATS | **2.46** | 5.58 | 1.44 | 1.81 | 4.77 |
| ARNN | 3.29 | 5.67 | 3.16 | 2.23 | 2.83 |
| ARIMA-ARNN | 3.59 | 5.50 | **0.71** | 2.06 | 1.53 |
| ARIMA-WARIMA | 3.64 | 5.69 | 0.72 | 2.05 | 1.58 |
| Proposed TARNN | 3.27 | **5.07** | 1.60 | **1.53** | **1.05** |
| RMSE values |  |  |  |  |  |
| ARIMA | 85378.21 | 19598.5 | 5325.44 | 2879.54 | 28661.69 |
| Theta | 77538.27 | 21033.15 | 9159.85 | 2976.44 | 22950.91 |
| WARIMA | 116791.17 | 43298.22 | 25609.64 | 2764.52 | 28139.06 |
| ETS | 60473.05 | 20017.92 | 6527.45 | 2425.78 | 33242.02 |
| TBATS | **60472.43** | 19855.08 | 7262.95 | 2647.24 | 86202.46 |
| ARNN | 79694.81 | 16796.84 | 14859.34 | 3063.08 | 48693.24 |
| ARIMA-ARNN | 86626.36 | 19557.8 | **5323.38** | 2876.86 | 27825.87 |
| ARIMA-WARIMA | 86938.13 | 20249.21 | 5428.55 | 2845.54 | 28908.41 |
| Proposed TARNN | 78320.37 | **16559.04** | 7854.66 | **2323.95** | **20423.69** |
| MAE values |  |  |  |  |  |
| ARIMA | 72823.9 | 17672.58 | 2833.76 | 2390.47 | 24714.92 |
| Theta | 66491.15 | 18797.76 | 8024.95 | 2497.19 | 20853.47 |
| WARIMA | 101923.84 | 36890.4 | 22279.76 | 2069.01 | 24294.93 |
| ETS | 50537.67 | 17953.84 | 4972.05 | 1893.77 | 29506.42 |
| TBATS | **50533.39** | 17839.37 | 6091.65 | 2074.26 | 75834.12 |
| ARNN | 67409.69 | 14941.82 | 13389.13 | 2556.93 | 44939.99 |
| ARIMA-ARNN | 73690.5 | 17585.56 | **3023.91** | 2386.66 | 24259.25 |
| ARIMA-WARIMA | 74637.97 | 18187.05 | 3056.05 | 2353.5 | 25162.57 |
| Proposed TARNN | 66939.65 | **14770.08** | 5487.11 | **758.82** | **16701.18** |

**Figure 3.**
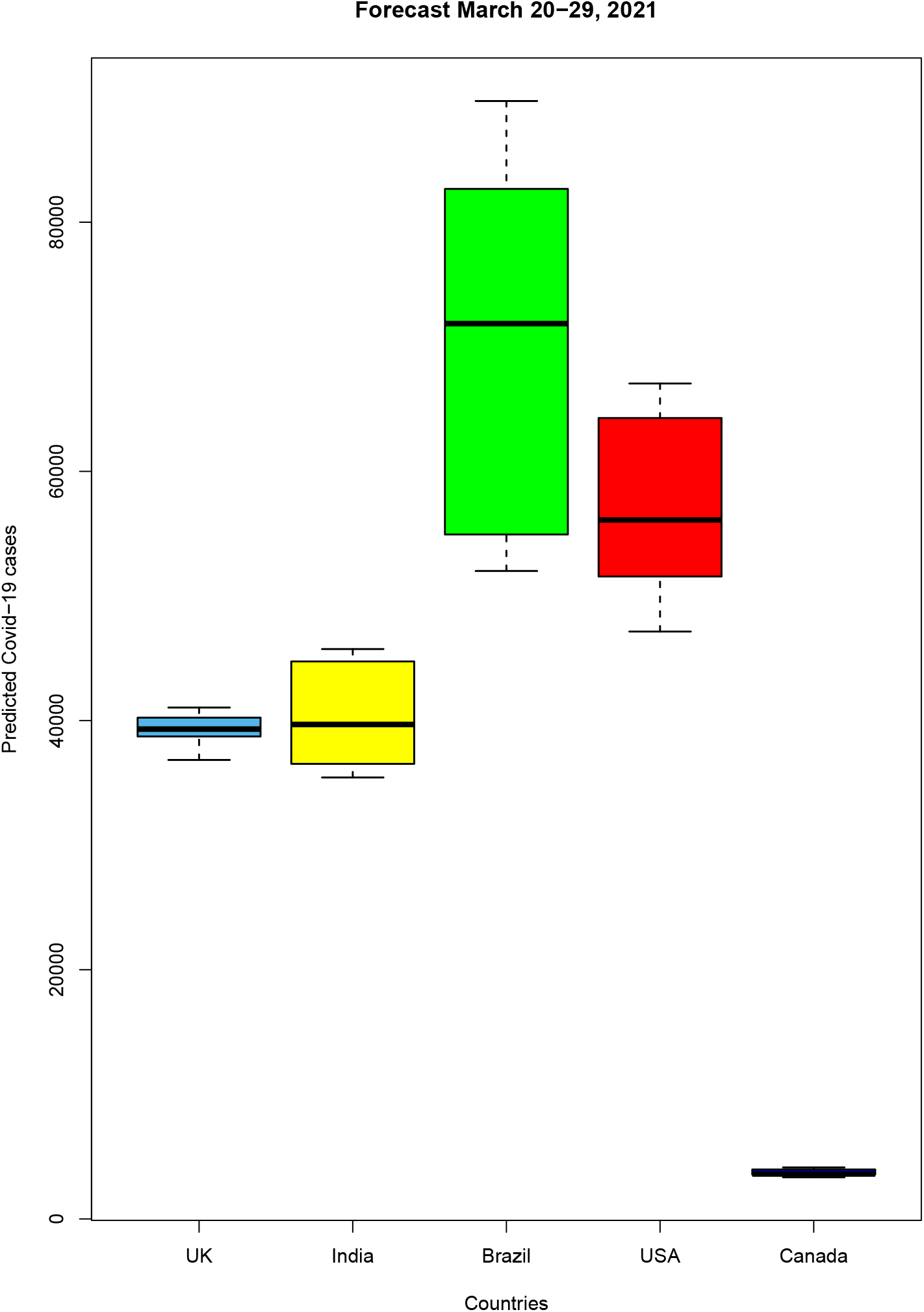
Forecast of COVID-19 cases for March, 2021 using proposed TARNN model.

## 5. Discussions and Conclusions

In this study, we proposed a novel hybrid Theta-ARNN (TARNN) model using residual modelling approach that performs considerably well for confirmed cases of COVID-19 forecasting for the countries that include the ones with the highest number of cases USA, followed by Brazil, India, UK, and Canada. The proposed TARNN model filters linearity using the Theta model and can better explain the linear, nonlinear, and nonstationary tendencies present in the selected COVID-19 data sets compared to the traditional single and hybrid models. It also yields better forecast accuracy than various traditional single and hybrid models for three out of five countries. The proposal will be useful in decision and policy making for government officials and policymakers to allocate adequate health care resources for the coming days in responding to the crisis. Time series of epidemics can oscillate heavily due to various epidemiological factors and these fluctuations are challenging to be captured adequately for precise forecasting. This newly developed model can still predict with better accuracy provided the conditions of asymptotic stationarity of the hybrid model are satisfied. This method can be used to update real-time forecasts as more data becomes available. The study covering multiple countries can be utilized without geographical borders and reflects the impact of social distancing, wearing masks, lock down, shutdown, quarantine, and sanitizing proper measures implemented by authorities. Prevalent techniques in literature were unable to completely capture the nonlinear behavior of stochastic time series containing inherent random shock components. This new method have significant theoretical (established ergodicity and stationarity of the proposed TARNN process) as well as practical implications. Authorities and health care can modify their planning in stockpiles and hospital-beds depending on these forecasts of the COVID-19 pandemic.

Many parameters associated with COVID-19 transmission are still poorly understood. The resulting model uncertainty is not always calculated or reported in a standardized way. Once we can incorporate these variables, we can improve our estimates and update the TARNN model accordingly. Since purely statistical approaches do not account for how transmission occurs, they are generally not well suited for long-term predictions about epidemiological dynamics (such as when the peak will occur and whether a resurgence will happen) or for inferences about intervention efficacy. Most forecasting models therefore limit their projections to one week or a few weeks ahead. Moreover, the problem of using confirmed cases to fit models is further complicated by the fact that the fraction of cases that are confirmed is spatially heterogeneous and time-varying. Amid enormous uncertainty about the future of the COVID-19 pandemic, the proposed TARNN model yields quantitative projections that policymakers may need in the short term to allocate resources or plan interventions. To conclude, this model can further be easily extended for similar nonlinear and non-Gaussian forecasting problems arising in other applied domains.

## Data Availability

Data and codes are available at: https://github.com/arinjita9/COVID-19-Forecasting-by-TARNN-

## Data and Code

All the results can be effortlessly updated with the help of R-shiny application as new data becomes available. This publicly available repository link https://github.com/arinjita9/COVID-19-Forecasting-by-TARNN- contains the current data files and R scripts for the TARNN model which ensures the repeatability and reproducibility of the results presented in this study.

